# A comprehensive genomic framework for identifying genes predisposing to homologous recombination repair deficient breast cancer

**DOI:** 10.1101/2025.10.22.25338181

**Authors:** José Camacho-Valenzuela, Thibaut Matis, Carla Roca, Jorge Luis Cuamatzi Flores, Nancy Hamel, Barbara Rivera, Simon Gravel, Paz Polak, Carla Daniela Robles-Espinoza, William D. Foulkes

## Abstract

**Background:** Patients with clinical characteristics of increased cancer susceptibility without an identified genetic lesion are regularly seen in clinics. Case-control studies and matched normal/tumour sequencing have advanced the discovery of Cancer Susceptibility Genes (CSGs), with limitations when used independently. We reasoned that combining these strategies alongside mutational signatures and clinical data could improve CSGs identification.

**Methods:** Using breast cancer exome data from The Cancer Genome Atlas (TCGA-BRCA), we developed a genomic framework that evaluates exome-wide associations of Germline Pathogenic Variants (GPVs) with somatic second hits, within the context of the Homologous Recombination Repair Deficiency (HRD) mutational signature 3 (Sig3). This is complemented by clinico-genomic analysis evaluating clinical and biological plausibility.

**Results:** Our framework confirmed significant associations with Sig3 of *BRCA1/2* GPVs with second hits, validating its performance. *THBS4* also reached significance but co-occurred with other HRD-related events. Borderline significance was observed for *KIF13B* and *TESPA1*. The clinico-genomics approach further identified *KIF13B* and *TESPA1*, as well as *RAD51B* and other Fanconi Anemia pathway-related genes, which deserve further validation.

**Conclusions:** Our framework strengthens identification of candidate HRD-related breast CSGs through combined statistical and clinico-genomics analyses. It is adaptable to other mutational signatures/cancer types and will benefit from larger and well-annotated datasets.

## Introduction

It has been observed that approximately 7-8% of cancer cases overall are attributable to inherited factors such as Germline Pathogenic Variants (GPVs) in Cancer Susceptibility Genes (CSGs)^1,2^. However, patients with clinical characteristics suggestive of increased cancer susceptibility that remain without identified GPVs in known CSGs are still regularly seen in the clinical setting, limiting options for genetic diagnosis, preventive strategies and treatment. This phenomenon, often known as “the missing heritability”, could be explained in part by as-yet undiscovered GPVs in unrecognized cancer risk genes^3^. Incorporating these missing genes in cancer susceptibility clinical gene panels could improve clinical management of patients.

Different high-throughput sequencing methods have advanced our understanding of genetic susceptibility to cancer, including case-control approaches^4–8^ and analysis of matched normal/tumour sequencing data^9–11^. Each approach has specific advantages and limitations. Case-control studies are powerful to assess statistically significant gene associations across large groups of patients, but they do not consider the biological context of each tumour and generally limit their analysis to predefined gene lists^6,7^. Meanwhile, in-depth study of the tumour mutational context of GPVs yields valuable biological insights^9,10,12–14^, but such studies historically lacked statistical power due to small cohort size, though this is changing^11^. Combining both approaches, together with clinical data and mutational signatures into a unified framework, could theoretically enable the identification of CSGs that may have remained undetected.

The matched normal/tumour exome dataset of breast cancer patients from The Cancer Genome Atlas (TCGA) is an extensively curated source of genomic and clinical data necessary for such an integrated analytical approach. Furthermore, the COSMIC (v3.4) Single-Base Substitution Signature 3 (Sig3) associated with Homologous Recombination Repair Deficiency (HRD) has a high prevalence in breast cancer^12,15,16^. Sig3 and HRD are associated with the Triple-Negative Breast Cancer clinical subtype^13,17^, which overlaps in approximately 80% of cases with Basal-like Breast Cancer (BLBC)^18^, one of the 5 intrinsic subtypes determined by the PAM50 classification^19,20^. Mechanistically, Sig3 typically results from biallelic inactivation of the well-known breast cancer risk genes *BARD1, BRCA1/2, PALB2,* and *RAD51C/D*^9,12–14,21^. As such, presence of Sig3 in a tumour could be used to infer the underlying biallelic inactivation of any gene (whether known or unknown) contributing to HRD^22^.

In this study, we developed a comprehensive genomic framework that integrates association analysis, matched normal/tumour exome data, and clinical information from the TCGA breast cancer cohort (TCGA-BRCA). The framework aims to identify GPVs with somatic second hits in patients where the presence of Sig3 in the tumour is not explained by genetic events in known HRD genes. Building on prior work^9,10,13,14,23,24^, we separately assessed the statistical association of GPVs with somatic second hits in breast cancer patients with and without Sig3, unrestricted to predefined gene sets. We integrated outputs from this dual statistical and patient-specific clinico-genomics approach to identify new candidate HRD-related breast CSGs.

## Methods

### Cohort data

We used exome data from the TCGA-BRCA cohort (dbGAP Project #34072: phs000178.v11.p8). Blood-derived BAM files were downloaded from the Genomics Data Commons (GDC) platform. Somatic mutation data was obtained from the Multi-Center Mutation Calling in Multiple Cancers (MC3) project^25^. Copy Number Variants (CNVs) data was obtained from the GDC platform. Somatic promoter methylation data for *BRCA1* and *RAD51C* was obtained from a previous study^13^, where they were analyzed for correlation with messenger RNA (mRNA) downregulation. Clinical information was obtained from the cBioPortal. A summary of the datasets and sources is provided in **Supplementary Table 1**. Further details are described in **Supplementary Methods**.

### Sample inclusion criteria

Sig3 detection relies on assessing single nucleotide somatic mutations in a tumour sample. To ensure reliable detection of Sig3, we established somatic mutation count thresholds. By performing bootstrap simulations in 14 positive controls downloaded from the COSMIC database (https://cancer.sanger.ac.uk/signatures/sbs/sbs3/), we determined a minimum threshold of 28 somatic mutations. On the other hand, to eliminate hypermutated tumours, we considered 500 somatic mutations as the maximum threshold, as reported by Van Marcke et al., 2020^24^. Selecting tumours with > 28 and < 500 somatic mutations from our cohort resulted in 824 patients eligible for the subsequent analyses.

### Bioinformatics pipeline for germline data

Blood-derived BAM files were processed using an in-house bioinformatics pipeline, and downstream exome filtering was performed according to predefined criteria (detailed in **Supplementary Methods**). Briefly, exome filtering included the following steps: **1)** Variants flagged in gnomAD v2.1.1 as failing at least one of their quality controls were excluded; **2)** Variants with >4 alternate reads, >20 total depth, and Variant Allele Fraction >0.1 were kept; **3)** Exonic and splice site variants were retained, specifically missense, nonsense, frameshift INDELs, start-loss, and stop-loss; **4)** Variants with population allele frequency < 0.01 in ExAC and/or gnomAD (or absent from both) were kept; **5)** Variants classified in ClinVar as pathogenic or likely pathogenic were retained; **6)** For variants annotated in ClinVar as conflicting, Variants of Uncertain Significance (VUS), or lacking classification, only missense, nonsense, splicing, and frameshift INDELs were further evaluated with stringent *in silico* predictions; **7)** Genes with 3 or more variants in the same patient were excluded to reduce potential residual noise; **8)** Olfactory receptor genes were also discarded due to false positive associations in cancer analyses^26^. The resulting final list of germline variants were considered as GPVs.

### Second hit and Sig3 analyses

We determined the somatic second hit status of each GPV per patient in the cohort, either as **1)** somatic point mutations/small INDELs or **2)** as Loss of Heterozygosity (LOH) through the loss of the wild type allele (further details in **Supplementary Methods**). From the 824 cases, 30 had missing/unavailable CNV data and were therefore excluded. This resulted in a total final cohort of 794 patients that were included in all subsequent analyses. We analyzed Sig3 presence for each individual patient’s tumour (n = 794) using SigMA^27^, a computational algorithm that accurately detects Sig3 in tumour samples with low mutation count. Utilizing the somatic mutation data of the MC3 project, a Sig3 score was calculated by the gradient boosting classifier of the algorithm for each tumour.

### Unbiased association analysis approach

We considered a tumour as “Sig3 positive” when having a Sig3 score >=0.5, as suggested by Gulhan et al., 2019^27^. Based on Sig3 scores, we divided the cohort (n = 794) in 2 subgroups: patients with Sig3 in their tumours (Sig3+ group) and patients without (Sig3-group), resulting in 141 and 653 patients, respectively. We calculated the frequency of carriers of GPVs with second hits in each gene of the Sig3+ group compared to their respective frequency of carriers of GPVs with second hits in the Sig3-group, as shown in **Figure 1**. We followed an unbiased exome-wide approach where we did not restrict the analysis to any gene panel. In addition, to better detect signals from potentially new genes beyond the well-known HRD genes, we also performed a sub-analysis excluding patients carrying GPVs in *BARD1, BRCA1, BRCA2, PALB2, RAD51C* and *RAD51D*.

**Figure 1.**
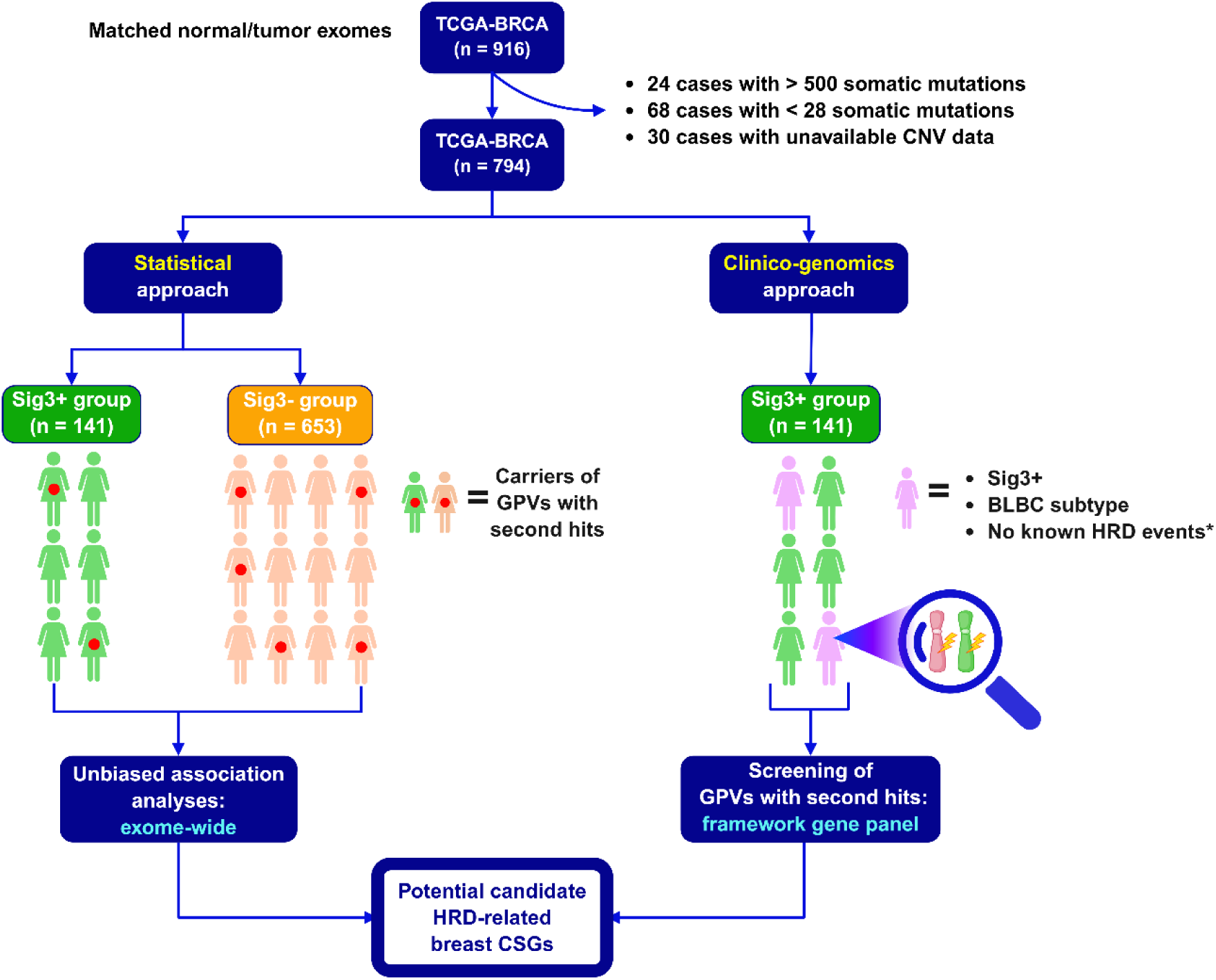
Schematic overview of the comprehensive genomic framework. Unbiased association analyses are shown on the left. The clinico-genomics approach is shown on the right. <u>Key</u>: *No known HRD events: 1) GPVs with second hits and somatic homozygous deletion in the 6 well-known HRD-associated genes *BARD1, BRCA1/2, PALB2* and *RAD51C/D*; 2) Somatic promoter methylation in *BRCA1* or *RAD51C*. Figure created with BioRender.com. <u>Abbreviations</u>: TCGA-BRCA: Breast cancer exome dataset of The Cancer Genome Atlas. CNV: Copy Number Variants. Sig3+: Signature 3 positive. Sig3-: Signature 3 negative. GPVs: Germline Pathogenic Variants. HRD: Homologous Recombination Repair Deficiency. BLBC: Basal-like Breast Cancer. CSGs: Cancer Susceptibility Genes.

### Statistical analysis and reproducibility

We performed comparisons for the unbiased association analysis by evaluating, in a case-only design, whether the frequency of carriers of GPVs with second hits in each gene was enriched in Sig3+ breast cancer patients compared to Sig3-cases. We used the Barnard’s exact test, (two-sided, p < 0.05), which does not condition on fixed margins for the evaluation of differences between 2 proportions analyzed in 2×2 contingency tables^28^. Such properties make this test particularly well-suited for our small sample size dataset, conferring greater statistical power to evaluate associations in the context of low event counts. Odds ratios and 95% Confidence Intervals (CIs) were calculated for each gene assessed with the Wald test. Here, the OR is measuring the magnitude of the association between the gene in question (with a second hit) with breast cancer that is Sig3+ compared with breast cancer that is Sig3-, not the overall risk of breast cancer. For genes in the Sig3-group with no observed events, the Haldane-Anscombe correction (adding 0.5 to genes with zero counts) was applied. Multiple hypotheses testing was mitigated with the Benjamini-Hochberg method. Due to the relatively small sample size of our cohort (794 cases), a large number of genes (3685 out of 4522, 81%) had very low counts of GPVs with second hits (1 or 2 events), limiting the ability to estimate associations. Therefore, for tests contrasting Sig3+ and Sig3-patients, we only considered genes harboring GPVs with second hits in at least 5 different patients as a minimum threshold to increase statistical power and reduce the risk of false associations driven by very low count of events.

### Clinico-genomics approach

To complement the unbiased association analyses, we incorporated biological principles (e.g. Knudson’s two-hit hypothesis) into the analytical framework as an attempt to reconstruct the most likely tumorigenesis path for each tumour. We integrated germline, tumour and clinical data of all the individual patients from the Sig3+ group (n = 141). We annotated validated types of genetic events that can explain the Sig3+ status of each tumour: both GPVs with second hits and somatic homozygous deletion in the 6 well-known HRD-associated genes *BARD1, BRCA1/2, PALB2* and *RAD51C/D*, as well as somatic promoter methylation in *BRCA1* or *RAD51C*. The rationale is that GPVs that co-exist with other genetic events already known to be linked to Sig3+ status in tumours are less likely to be viable candidates than GPVs in patients where no other putative drivers of Sig3+ status are observed.

We first looked for Sig3 patients with BLBC but lacking any annotated HRD-associated events that could explain the presence of Sig3. In these patients, we then identified GPVs that have second hits in the tumour from an expanded cancer-susceptibility gene panel (hereafter referred to as ‘framework gene panel’), a gene list curated in-house and created by merging clinical-grade panels for cancer susceptibility offered by different diagnostic laboratories, as well as genes evaluated in two of the largest case-control studies in hereditary breast cancer^6,7^, and a broad list of DNA-repair genes (**Supplementary Data 1**). The inclusion of DNA-repair genes is based on the hypothesis that, in addition to *BARD1, BRCA1/2, PALB2* and *RAD51C/D*, other homologous recombination repair genes currently not known to be related to Sig3 (or genes from other DNA-repair pathways) may also contribute to HRD. We also included the genes reaching statistical significance in the exome-wide unbiased association analyses.

## Results

### Identifying candidate CSGs using unbiased association analyses

In **Figure 1**, we illustrate a schematic overview of the comprehensive genomic framework proposed in our study. The unbiased association analysis compares the per gene frequency of carriers of GPVs with second hits between Sig3+ and Sig-patients. In a first, global association analysis (without exclusions), a total of 221 genes were assessed. We observed a significant association (p < 0.05) of Sig3 with presence of GPVs with second hits in the proof of principle genes *BRCA1* with the highest odds ratios (OR = 38.7 [95% CI 8.7 – 171.5], adjusted p-value 1.8×10^-8^) and *BRCA2* (OR = 30.2 [95% CI 6.6 –136.9], adjusted p-value 2.06×10^-6^), as illustrated in **Figure 2** and **Supplementary Data 2**. This observation was expected and validates the effectiveness of the framework. In this case-only design, the odds ratios indicate that carriers of *BRCA1/2* GPVs with second hits are >30 times more likely to be Sig3+ than Sig3-. We also detected a significant association (p < 0.05) of *THBS4* with high odds ratios (OR = 23.9 [95% CI 2.7 – 206.8], adjusted p-value 0.02), indicating enrichment in Sig3+ breast cancer patients over Sig3-cases. No other gene reached statistical significance (p < 0.05) for association with presence of Sig3. Notably, some well-known HRD genes were not statistically tested because they did not meet the minimum threshold of harboring GPVs with second hits in at least n = 5 different patients to qualify for statistical comparison; this is the case for *BARD1* (n = 1), *PALB2* (n = 2), *RAD51C* (n = 2) and *RAD51D* (n = 1). Interestingly, other breast CSGs commonly tested clinically (*ATM* and *CHEK2*) showed odds ratios below 1 (**Figure 2** and **Supplementary Data 2**) which, while not statistically significant, were suggestive of a negative association with Sig3 (*ATM:* OR = 0.2 [95% CI 0.03 – 2.1], adjusted p-value 0.54; *CHEK2*: OR = 0.3 [95% CI 0.01 – 5.3], adjusted p-value 0.58).

**Figure 2.**
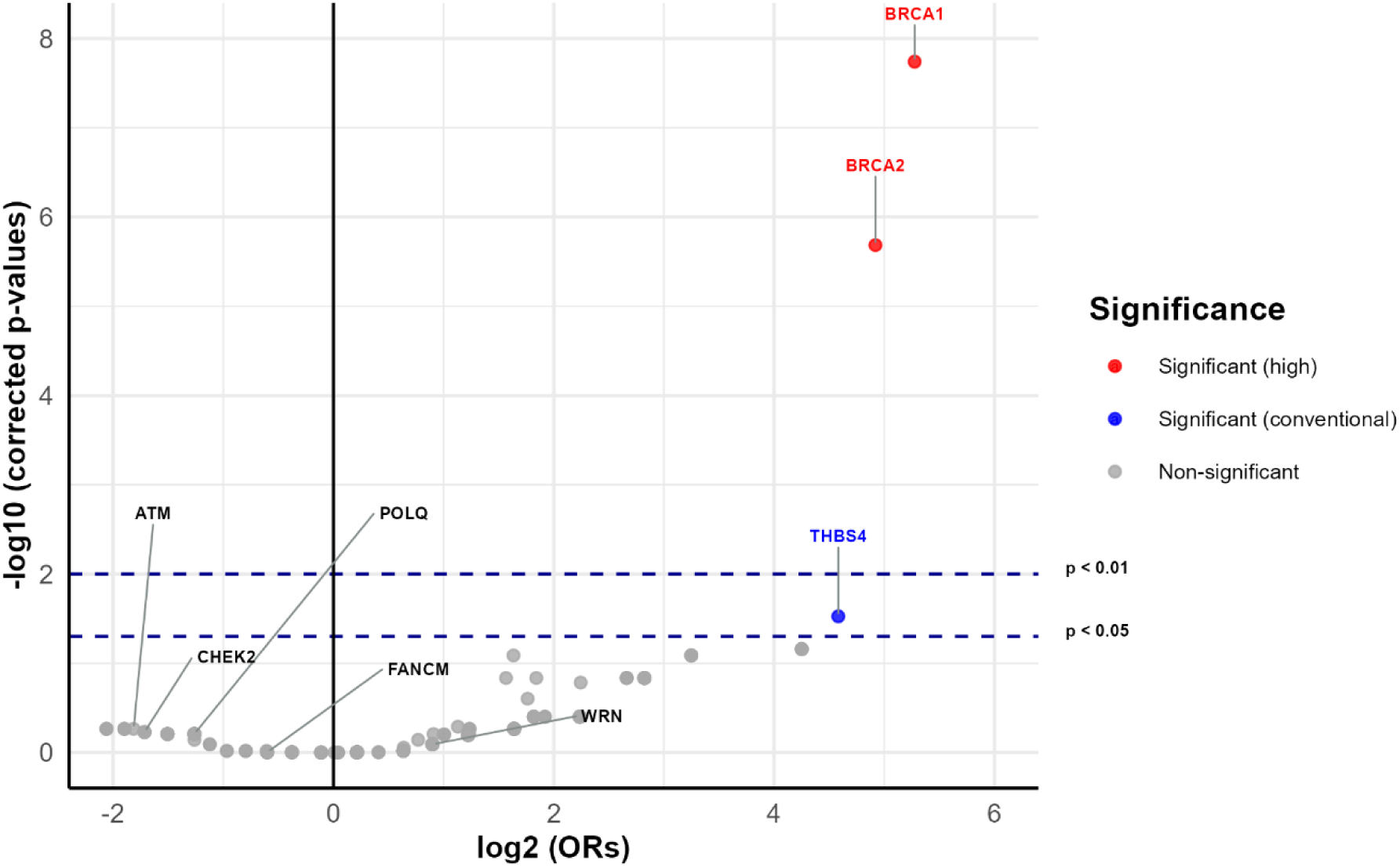
Exome-wide association with Sig3 of GPVs with somatic second hits, which included patients carrying GPVs with second hits in the well-known HRD genes *BARD1, BRCA1/2, PALB2,* and *RAD51C/D*. Barnard’s exact test, two-sided. The X axis shows odds ratios (log2 scale). The Y axis shows adjusted p-values (-log10 scale). Horizontal dotted dark-blue lines indicate the conventional (p < 0.05) and strict (p < 0.01) significance thresholds. P-values shown were adjusted with the Benjamini-Hochberg method to mitigate multiple hypotheses testing. Red dots indicate genes reaching statistical significance at p < 0.01. The blue dot indicates the gene that reached statistical significance at p < 0.05. Gray dots indicate genes not reaching statistical significance. Non-significant genes labeled in the plot correspond to those included in our framework gene panel. <u>Abbreviations</u>: Odds ratios: ORs.

To better detect signals from potential new genes beyond the already well-known HRD genes, we repeated the unbiased exome-wide association analysis by excluding all the patients with GPVs with second hits in the well-known HRD genes *BARD1, BRCA1*, *BRCA2, RAD51C, RAD51D* and *PALB2*, resulting in 194 genes being analyzed. Here, *THBS4* was again found to be statistically significant (OR = 24.8 [95% CI 2.7 – 224.4], adjusted p-value 0.04), as shown in **Figure 3** and **Supplementary Data 3**. In addition, in the context of fewer gene comparisons tested, 3 additional genes emerged as statistically significant: *KIF13B* and *TESPA1* scored the same association numbers as *THBS4* since they all had the same number of events in both Sig3+ group (n = 4) and Sig3-group (n = 1), and *ATXN3* had a more modest odds ratios (OR = 4.3 [95% CI 2.04 – 9.3], adjusted p-value 0.04). In the case of *ATXN3*, most of the identified variants in both Sig3+ and Sig3-groups were frameshift insertions clustered within a low complexity genomic region flagged in gnomAD as “lcr” (except for two variants which were located only 2 and 3 base pairs outside this area, respectively). Such regions are characterized by repetitive sequences prone to alignment issues and sequencing artifacts. While we retained this gene in the association analyses to preserve an unbiased exome-wide approach, its biological interpretation is likely artifactual. No other gene reached statistical significance (p < 0.05). For *ATM* and *CHEK2*, removing the Sig3+ cases already attributable to known HRD-associated genes resulted in ORs that were as expected slightly closer to 1.0 (*ATM* (OR = 0.36 [95% CI 0.04 – 2.8], adjusted p-value 0.74) and *CHEK2* (OR = 0.39 [95% CI 0.02 – 6.9], adjusted p-value 0.74).

**Figure 3.**
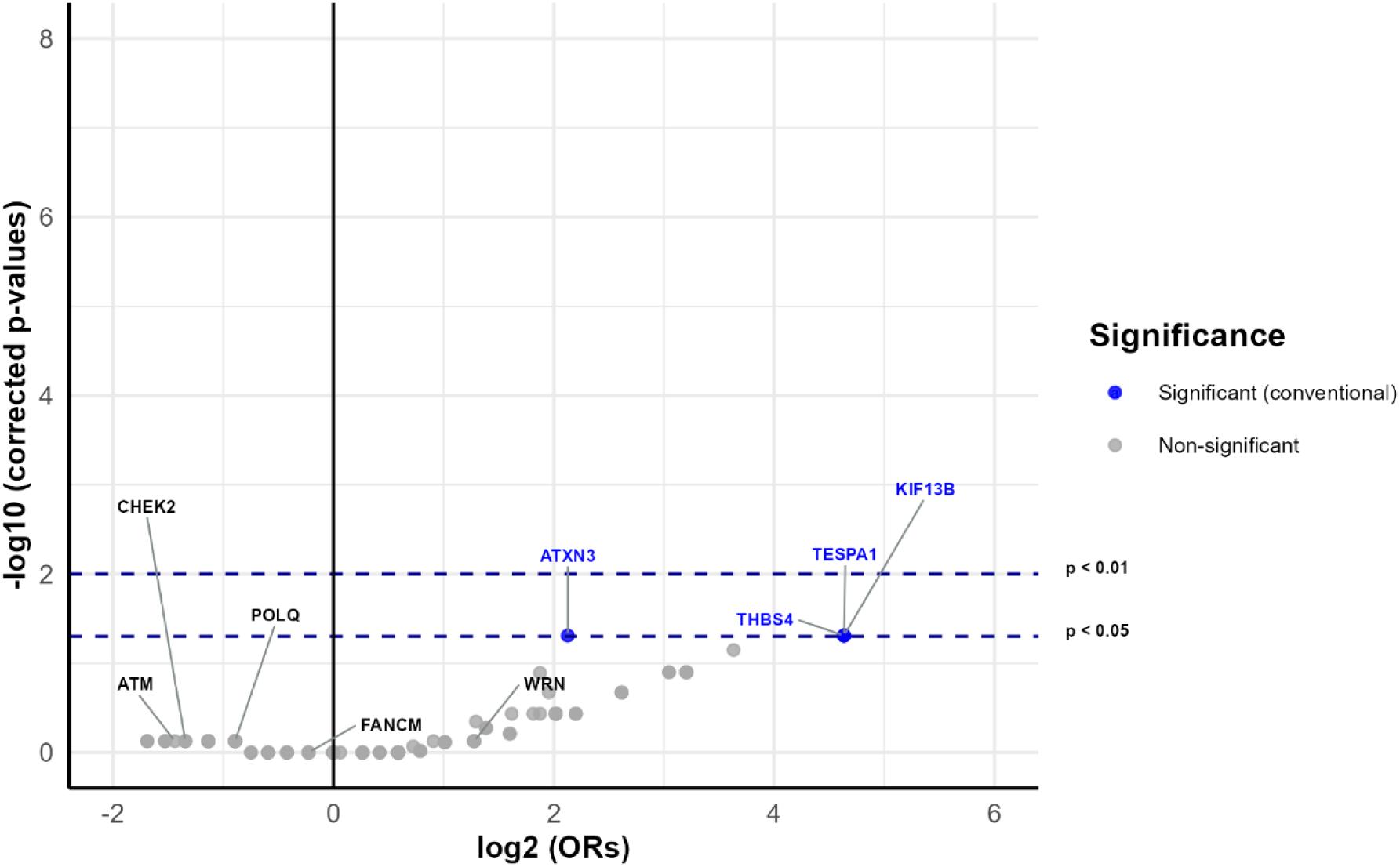
Exome-wide association with Sig3 of GPVs with somatic second hits, which excluded patients carrying GPVs with second hits in the well-known HRD genes *BARD1, BRCA1/2, PALB2,* and *RAD51C/D*. Barnard’s exact test, two-sided. The X axis shows odds ratios (log2 scale). The Y axis shows adjusted p-values (-log10 scale). Horizontal dotted dark-blue lines indicate the conventional (p < 0.05) and strict (p < 0.01) significance thresholds. P-values shown were adjusted with the Benjamini-Hochberg method to mitigate multiple hypotheses testing. Blue dots indicate genes that reached statistical significance at p < 0.05. Gray dots indicate genes not reaching statistical significance. Non-significant genes labeled in the plot correspond to those included in our framework gene panel. <u>Abbreviations</u>: Odds ratios: ORs.

### Identifying candidate CSGs using a clinico-genomics approach

We jointly evaluated the clinical data (intrinsic clinical subtype, age at diagnosis, sex, and TCGA-reported race) of each individual patient of the Sig3+ group (n = 141) with their germline and somatic mutational landscape (**Supplementary** Figure 1). We screened GPVs with second hits in genes from our in-house framework gene panel in Sig3+ patients with BLBC that lacked any well-known HRD-associated events that could explain the presence of Sig3 in their tumours. We included in our framework panel the statistically significant genes identified by our unbiased association analyses, *KIF13B, TESPA1* and *THBS4*.

Using this strategy, we identified 11 of 141 patients from the Sig3+ group (7%) that carried 17 GPVs with second hits in 16 different genes (**Figure 4** and **Supplementary Data 4**). Of these 11 patients, 6 cases harbored 6 GPVs with second hits in 5 known CSGs, including *ALK, BLM, FANCM, RET* and *SMARCE1*, all of which were missenses. Likewise, 9 patients carried 12 GPVs with second hits in 11 DNA-repair genes, including *ATR, BLM, DCLRE1A, ERCC4, FAAP20, FAN1, FANCD2, FANCM, LIG3, PARP3* and *RAD51B* (**Supplementary Data 4**). Of note, 6 of 12 GPVs with second hits were predicted to be truncating, including 3 nonsenses in *ATR, PARP3* and *RAD51B*, 1 frameshift deletion in *FAAP20*, and 2 splicing in *FAN1* and *FANCD2*.

**Figure 4.**
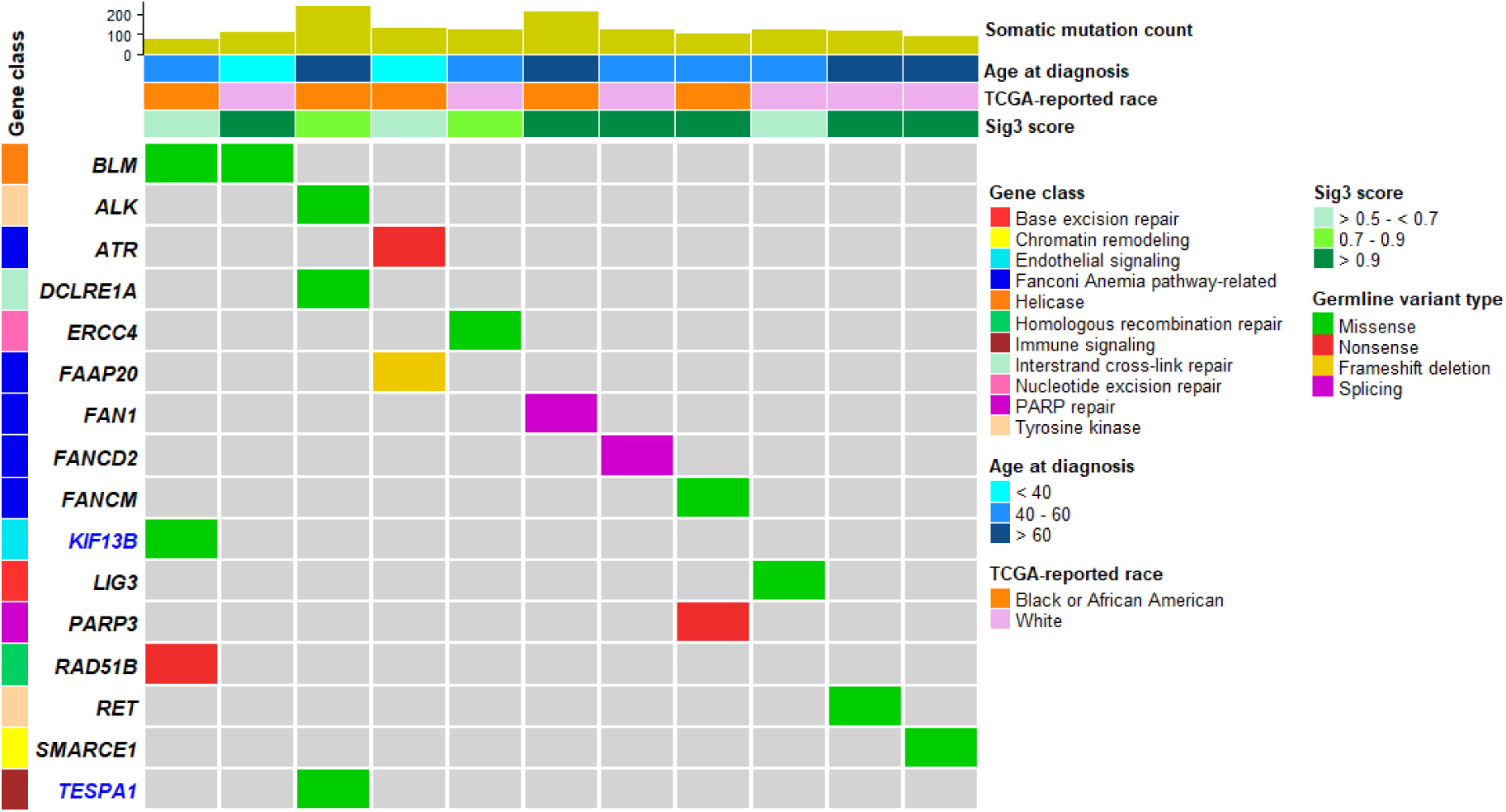
Germline and somatic mutational landscape of Sig3+ patients (n = 11/141) with BLBC identified through the clinico-genomics approach in the TCGA-BRCA cohort. All the GPVs illustrated harbored a second hit in the tumour, and no HRD-related genetic events (GPVs with second hits and somatic homozygous deletions in *BARD1, BRCA1/2, PALB2,* and *PALB2*, as well as somatic promoter methylation in *BRCA1/RAD51C*) were observed in such patients. All patients had the BLBC intrinsic subtype. Gene labels in blue indicated genes that also reached statistical significance in the unbiased exome-wide association analyses (p < 0.05). <u>Abbreviations</u>: TCGA-BRCA: Breast cancer exome dataset of The Cancer Genome Atlas. Sig3: Signature 3. PARP repair: poly (ADP-ribose) repair.

To get a global view of the Sig3+ group, we also examined Sig3+ patients with well-known HRD-related events that could explain the presence of the mutational signature. Specifically, we identified 73/141 patients (51%) carrying a well-known HRD-related event, of whom 72/73 carried a single HRD-related event, distributed as follows (**Figure 5**): 38 cases with a *BRCA1*-related HRD event (15/38 with *BRCA1* GPVs with second hits and 23/38 with *BRCA1* somatic promoter methylation), 15 cases with a *BRCA2*-related HRD event (12/15 with *BRCA2* GPVs with second hits and 3/15 with *BRCA2* somatic homozygous deletion), 15 cases with a *RAD51C*-related event (2/15 with *RAD51C* GPVs with second hits, 12/15 with *RAD51C* somatic promoter methylation and 1/15 with *RAD51C* somatic homozygous deletion), 2 cases with a *PALB2*-related HRD event (2 *PALB2* GPVs with second hits), 1 case with a *RAD51D*-related HRD event (a *RAD51D* GPV with second hit), and 1 case with a *BARD1*-related HRD event (a *BARD1* GPV with second hit). The remaining patient (1/73) had co-occurrence of 2 different HRD-related events, namely somatic promoter methylation in both *BRCA1* and *RAD51C*.

**Figure 5.**
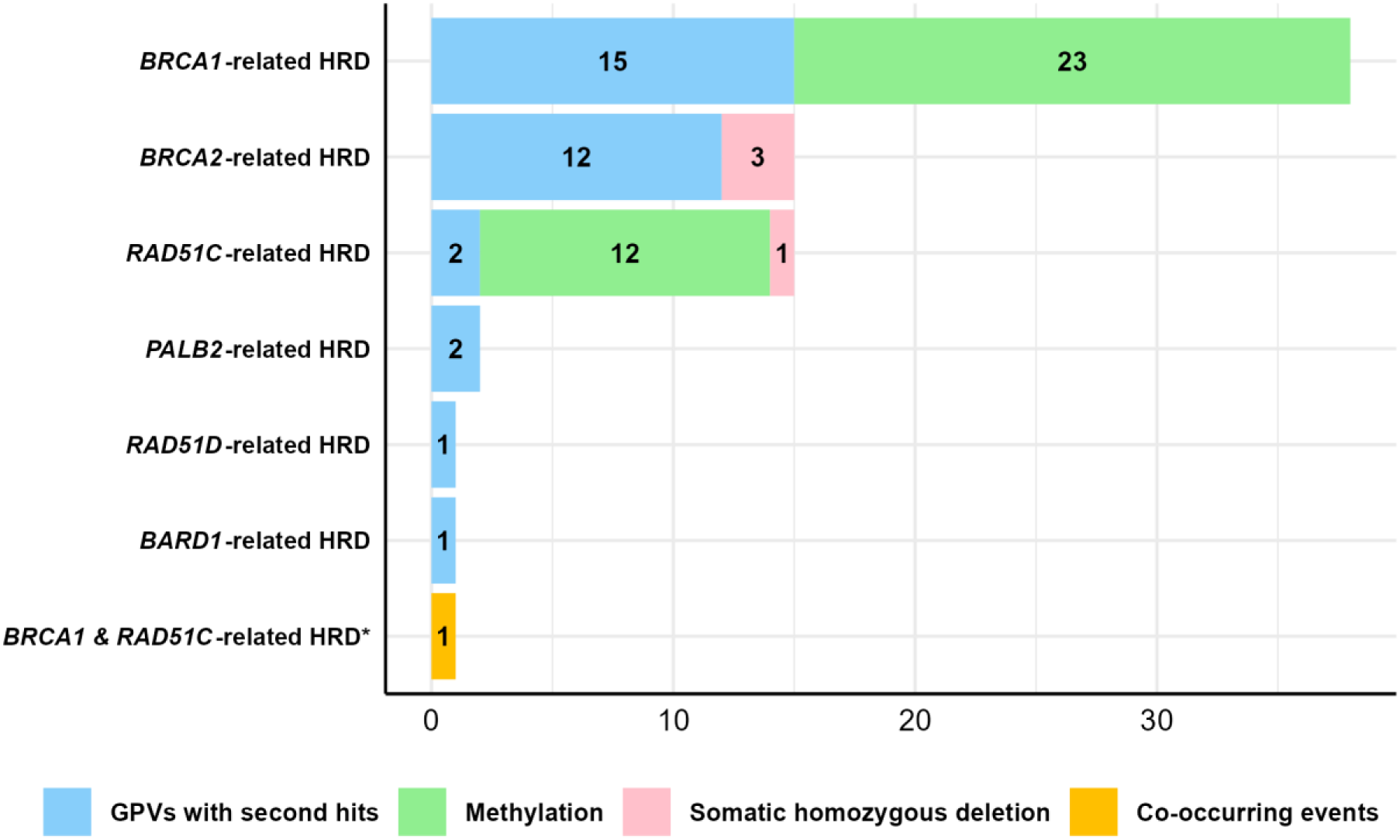
Sig3+ patients (n = 73/141) with well-known HRD-related genetic events that potentially explains the presence of the mutational signature in question. <u>Key</u>: \**BRCA1 & RAD51C*-related HRD: Patient with somatic promoter methylation in both *BRCA1* and *RAD51C*. <u>Abbreviations</u>: HRD: Homologous Recombination Repair Deficiency. GPVs: Germline Pathogenic Variants.

## Discussion

In this study, we developed and optimized a comprehensive genomic framework designed to investigate cancers where mutational signatures in the tumour can be leveraged to identify CSGs in the germline, with the goal to explain part of the missing heritability still observed in cancer genetics clinics. Our framework integrates a statistical unbiased association analysis of GPVs with their second hit tumour status within the context of cancer-relevant mutational signatures. This first analysis is then complemented by clinico-genomics sequencing evidence to provide both biological and clinical plausibility for the statistical findings.

We built on established matched normal/tumour sequencing frameworks from previous studies^9,10,13,24^ to integrate the somatic mutational landscape of each GPVs carrier, adding an additional layer of control for co-occurring events linked to the mutational signature under evaluation (here Sig3). This tailored approach allows us to attribute with higher confidence GPVs with second hits in candidate genes to the mutational signature under analysis, all within the context of the clinical subtype associated with such signature.

Our framework is also versatile. It can be used investigate other clinically relevant mutational signatures and can be extended to different cancer types associated to a specific mutational process, (e.g. mismatch repair signatures in colorectal cancer). Here, we used the TCGA-BRCA cohort as proof of principle for the framework because the dataset contains multiple levels of well-curated genomic and clinical data, and Sig3’s association with breast CSGs is well established. However, this framework will perform best to identify novel CSGs when applied to a larger cohort to provide sufficient statistical power to capture rare events and to distinguish true associations from false positives.

### Statistically significant CSGs identified by association analyses

The framework’s unbiased exome-wide analysis identified the expected proof of principle genes *BRCA1/2* with the highest statistical significance (**Figure 2**). The very high odds ratios observed for *BRCA1/2* reflect the case-only design of our analysis (Sig+ vs Sig3-in breast cancer patients) rather than case-control risk. For *BRCA1*, they also likely capture a BLBC-specific signal, since 14/15 carriers of *BRCA1* GPVs with second hits were BLBC. Similar odds ratios were reported in the CARRIERS study, where GPVs in *BRCA1* were strongly associated with TNBC^6^. Additionally, *THBS4* reached statistical significance (p < 0.05) in both association analyses (with and without inclusion of carriers of GPVs with second hits in the well-known HRD genes). *THBS4* is an extracellular matrix gene previously suggested to be possibly a tumour suppressor^29^, implicated in breast cancer^30^, and observed in hepatocellular and gastric cancers^31,32^. However, the clinico-genomics assessment showed that *THBS4* GPVs with second hits co-occurred either with other HRD-related events that better explain the presence of Sig3 in such carriers or were present in tumours with a non-BLBC clinical subtype (**Supplementary** Figure 1).

*KIF13B* (involved in VEGFR2 trafficking and angiogenesis^33^) and *TESPA1* (involved in the regulation of antitumour immunity^34^) neared statistical significance (p < 0.05) in the global unbiased association analysis and reached significance when carriers of GPVs (with second hits) in known HRD-related genes were removed from the analysis. In contrast with *THBS4,* after evaluating them in their respective clinico-genomics context, GPVs with second hits in *KIF13B* and *TESPA1* were observed in Sig3 patients with BLBC without HRD-related events in *BARD1*, *BRCA1/2*, *PALB2* or *RAD51C/D*. For the *KIF13B* carrier, the co-occurrence of a *RAD51B* GPV with second hit provides a more plausible explanation of the observed Sig3 given the role of *RAD51B* in homologous recombination repair. In contrast, for the *TESPA1* carrier, no alternative genomic or promoter methylation event was found to explain the Sig3+ status beyond the *TESPA1* GPV with second hit. Despite its biological function not been linked to homologous recombination repair^35^, further investigation could be warranted for *TESPA1*.

### CSGs identified through the clinico-genomics approach

The clinico-genomics approach of our framework identified additional candidates, including *RAD51B* and the Fanconi Anemia pathway-related genes *ATR, FAAP20, FAN1, FANCD2,* and *FANCM*, most of which could not be statistically tested due to low event counts (except for *FANCM*). As a paralog of *RAD51C*/*D* (both established breast CSGs associated with HRD), *RAD51B* also promotes DNA double-strand break repair via homologous recombination^36,37^. Its candidacy as a plausible HRD-related breast CSG is supported by an external, TCGA-independent case-control study reporting a statistically significant association of *RAD51B* with breast and ovarian cancer^38^. Similarly, the 4 carriers of GPVs with second hits in *ATR, FAAP20, FAN1, FANCD2* and *FANCM* (of which 1 patient carried a GPV with second hit in both *ATR* and *FAAP20*) shared notable features: 3 out of 4 cases were diagnosed before 50 years, and 3 out of 4 were reported as Black or African American. All 5 genes functionally interact with the Fanconi Anemia pathway^39^, a DNA-repair mechanism traditionally linked to hereditary breast cancer and biologically related to the homologous recombination repair pathway^40,41^. GPVs in *ATR*^42,43^ and *FAN1*^44,45^ have been observed in hereditary breast cancer cases, and a case-control study found a statistically significant association of *FANCD2* with this disease^46^. *FAAP20* has been found to play a strong role in homologous recombination repair^47^.

Of interest, while we observed a suggestive but non-significant (p < 0.05) negative association of *FANCM* with Sig3 in our statistical analyses, our clinico-genomics approach identified this gene as a candidate susceptibility gene with a second hit in a Black or African American Sig3+ case with BLBC that lacked any known HRD events (**Figure 4**).

However, this patient also carried a nonsense *PARP3* GPV with a somatic second hit, raising questions about the respective contribution of each gene, if any, to the HRD phenotype. In the Sig3-control group, 4/7 *FANCM* carriers of GPVs with second hits were also BLBC, and 2/7 were reported as Black or African American (**Supplementary Data 5**). These findings are consistent with prior studies that link *FANCM* with BLBC^48–50^, but they may also indicate that the contribution of *FANCM* GPVs to HRD may depend on additional gene-specific co-occurring variants and/or population-specific genetic factors. For instance, a recent study reported a strong association of *FANCM* with Estrogen Receptor negative (ER-) breast cancer in a Hispanic/Latino cohort^51^, an association not observed in two of the largest case-control studies conducted in predominantly European cohorts^6,7^.

### Comparison with previous studies of HRD and novel breast CSGs

Prior studies have combined matched normal/tumour sequencing data with Sig3 in predefined gene sets to investigate non-*BRCA1/2* familial breast cancer cases^9,10,14,24^, to directly associate Sig3 with candidate CSGs^13^, or to compare mutational signatures in non-*BRCA1/2* breast tumours harboring candidate gene alterations with mutational signatures from *BRCA1/2*-mutated tumours^23^. Our framework builds on these studies by requiring GPVs under study to have a somatic second hit to test their association with Sig3 exome-wide (beyond predefined gene sets). In addition, it incorporates a layer of control for co-occurring events extended to all the known HRD genes, while leveraging BLBC clinical subtype to guide the identification of candidate CSGs. Together, these methodological refinements yield a framework that preserves and build on the strength of the approaches discussed above, while also supporting more credible Sig3 attribution to candidate genes.

### Framework Strengths and Limitations

The findings generated by our framework showcase the power of combining multiple approaches to cross-validate candidate susceptibility genes. They also highlight the advantage of including datasets with multiple levels of annotation such as copy number and methylation analyses. For example, these results show that the number of patients with somatic promoter methylation in *BRCA1* and *RAD51C* (21 and 12, respectively) far exceeded the number of patients carrying a GPV with second hits in such genes (15 and 2, respectively). Similarly, for *BRCA2*, including patients with somatic homozygous deletion (n = 3) increased the number of *BRCA2*-related HRD events by ∼20% over *BRCA2* GPVs with second hits alone (n = 12). These results show that focusing exclusively on germline testing may underestimate HRD prevalence in breast cancer patients. Likewise, they also have direct clinical implications to guide treatment-informed decisions in clinics, potentially expanding breast cancer patients’ eligibility for targeted therapy with poly (ADP-ribose) polymerase (PARP) inhibitors. This feature of our framework is not limited to HRD. It can be adapted to annotate validated genomic events from other cancer-associated mutational processes, such as mismatch repair deficiency in colorectal or endometrial cancer.

Our framework’s approach also has limitations. First, the use of whole exome sequencing, while more accessible than whole genome sequencing, excludes non-coding genetic variants possibly contributing to cancer risk and HRD. Second, since our framework evaluates only GPVs with second hits, it may overlook monoallelic variants potentially acting through haploinsufficiency. Third, the framework requires very large sample sizes to be effective. The relatively small sample size of the TCGA-BRCA dataset limited the statistical power to evaluate associations, requiring the adoption of a threshold for considering genes with GPVs with second hits in at least 5 different patients to minimize the risk of false associations, which led to some genes not being analysed. Finally, although our framework was able to flag biologically plausible candidate CSGs, the predominance of European-ancestry patients in TCGA-BRCA restricted the discovery of potential ancestry-related GPVs^52^. Generating large and more ethnically diverse datasets with complete germline and tumour sequencing, methylation and copy number data that can unlock the full power of our framework will be resource intensive.

## Conclusions

In summary, we present in this study a comprehensive genomic framework that integrates multiple types of germline/tumour genetic events and clinical information to identify novel cancer susceptibility genes and alleles. The versatility of our framework allows for the investigation of multiple mutational signatures and cancer types associated with mutational signatures. The results of applying our framework to the TCGA-BRCA cohort support previous data suggesting that *RAD51B* is an HRD-related breast CSG. Our framework also flagged other interesting candidates related to the Fanconi Anemia pathway: *ATR, FAAP20, FAN1, FANCD2,* and *FANCM*. Among others, these genes deserve further study for validation, especially their inclusion in both larger and ancestrally diverse case-control studies and matched normal/tumour sequencing research efforts.

## Data Availability

Germline sequencing data (blood-derived BAM files) analyzed in this study are available via the Genomic Data Commons (TCGA-BRCA, dbGaP Project #34072: phs000178.v11.p8). Somatic mutation data was obtained from the Multi-Center Mutation Calling in Multiple Cancers (MC3) project. Copy number variation data was downloaded from the Genomic Data Commons. Somatic promoter methylation data for *BRCA1* and *RAD51C* was obtained from a previous study^13^. Clinical data was downloaded from the cBioPortal. Further details and access links are provided in **Supplementary Table 1** and **Supplementary Methods**. The code used to perform the statistical and clinico-genomics analyses from the genomic framework presented in this study is publicly available via GitHub (https://github.com/jcv333).

## Acknowledgements

We would like to thank Fiona Chan Pak Choon for her guidance with downstream exome analyses, Irving Simonin Wilmer for his guidance with statistical analyses in R, and Shelly Alkoby for her assistance with the conformation of the framework gene panel.

## Author contribution

J.C.V. developed the computational genomic framework and bioinformatics pipelines, analyzed the data, and drafted the manuscript. T.M., C.R., and J.L.C.F. contributed to the pipelines’ development. S.G., C.D.R.E., and P.P. contributed to the design of the statistical analyses. W.D.F., N.H., C.D.R.E., and P.P. designed the project. N.H. obtained access to controlled germline data from TCGA. P.P. provided curated promoter methylation data. W.D.F., N.H., C.D.R.E., P.P., and B.R. provided supervision and scientific guidance. W.D.F., N.H., and C.D.R.E. supervised all aspects of the project. J.C.V., W.D.F., N.H., and C.D.R.E. wrote the manuscript. All authors reviewed the manuscript.

## Ethics approval and consent to participate

The Cancer Genome Atlas (TCGA) consortium collected tumor and non-tumor biospecimens from participating human subjects with informed consent and under the authorization of the corresponding institutional review boards. Further information is available at https://cancergenome.nih.gov/abouttcga/policies/informedconsent. Access to controlled germline sequencing data (blood-derived BAM files) from TCGA was obtained through the database of Genotypes and Phenotypes (dbGaP) and the Genomic Data Commons (GDC) portal under an approved project (Project #34072; accession phs000178.v11.p8). Additional information regarding TCGA ethics and data use policies is available at: https://www.cancer.gov/ccg/research/genome-sequencing/tcga/history/ethics-policies

## Author information

Department of Human Genetics, McGill University, Montréal, QC, Canada

José Camacho-Valenzuela, Simon Gravel & William D. Foulkes

Cancer Research Program, Centre for Translational Biology, The Research Institute of the McGill University Health Centre, Montréal, QC, Canada

José Camacho-Valenzuela, Nancy Hamel & William D. Foulkes

Lady Davis Institute for Medical Research, Jewish General Hospital, Montréal, QC, Canada

José Camacho-Valenzuela & William D. Foulkes

Cancer Genetics Unit, Institut Bergonié, Bordeaux, France.

Thibaut Matis

BRIC (BoRdeaux Institute of onCology), UMR1312, INSERM, Univ. Bordeaux, F-33000, Bordeaux, France.

Thibaut Matis

Bellvitge Biomedical Research Institute (IDIBELL), Avinguda de la Granvia de l’Hospitalet 199, 08908 L’hospitalet de Llobregat, Barcelona, Spain

Carla Roca & Barbara Rivera

International Laboratory of Research of the Human Genome, National Autonomous University of Mexico, Querétaro, México

Jorge Luis Cuamatzi Flores & Carla Daniela Robles-Espinoza

Gerald Bronfman Department of Oncology, McGill University, Montréal, Quebec, H4A 3T2, Canada

Barbara Rivera

Quest Diagnostics, Secaucus, NJ 07094, USA

Paz Polak

Experimental Cancer Genetics Group, Wellcome Trust Sanger Institute, Hinxton, Cambridge, United Kingdom

Carla Daniela Robles-Espinoza

**Corresponding author** Correspondence to William D. Foulkes.

## Additional information

Correspondence should be addressed to William D. Foulkes

## Supplementary Information

Supplementary information provided in this study includes Supplementary Methods, Supplementary Figure 1, Supplementary Table 1, and Supplementary Data 1-5.

