## Supplementary Information for "A comprehensive genomic framework for identifying genes predisposing to homologous recombination repair deficient breast cancer"

### Supplementary Methods

#### *Cohort data*

We selected exome data from the TCGA breast cancer cohort (TCGA-BRCA, dbGAP Project #34072: phs000178.v11.p8). Blood-derived BAM files were downloaded from the Genomics Data Commons (GDC) platform (https://portal.gdc.cancer.gov/). We excluded cases lacking a matched blood-derived BAM file. Somatic mutation data was obtained from the MC3 project[1], which provides the publicly available Mutation Annotation Format (MAF) file mc3.v0.2.8.PUBLIC.maf.gz (https://gdc.cancer.gov/about-data/publications/mc3-2017) containing the somatic mutation of 1026 breast cancer cases. CNV data was obtained from the GDC website (https://gdc.cancer.gov/about-data/publications/pancanatlas), downloading the ABSOLUTE-annotated seg file analyzed with the computational algorithm ABSOLUTE[2]. Somatic promoter methylation data for *BRCA1* and *RAD51C* was obtained from a previous study[3], where they were analyzed for correlation with messenger mRNA downregulation. Clinical information was downloaded from the cBioPortal (https://www.cbioportal.org/study/clinicalData?id=brca_tcga_pan_can_atlas_2018).

#### *Bioinformatics pipeline for germline data*

Blood DNA-derived BAM files were realigned to the human reference genome hg19 with the Burrows-Wheeler Aligner (BWA) version 0.7.17[4]. PCR duplicates were marked and removed using MarkDuplicates (Picard) of The Genome Analysis Toolkit (GATK)[5] version 4.1.2.0. Variant calling was performed individually for each sample with HaplotypeCaller (GATK 4.1.2.0), with a minimum confidence threshold of 20. To allow a hard-filtering step on the germline calls, we used VariantFiltration (GATK 4.1.2.0) considering the following criteria: **1)** Quality by Depth (QD) > 3.0; **2)** Fisher Strand (FS) < 100.0; **3)** Mapping Quality (MQ) > 30.0. Germline variant calls that met such 3 criteria were flagged as "PASS". The resulting Variant Call Format (VCF) files (n = 824) were annotated individually with Annovar[6] via command-line utilizing multiple databases: ExAC[7], gnomAD v2.1.1[8], ClinVar (release 20240917)[9], dbNSFP v4 (dbnsfp47a)[10], dbscSNV v1.1 (dbscsnv11)[11], dbSNP 151 (avsnp151)[12] and refGene (UCSC Genome Browser)[13].

Downstream exome filtering was carried out according to the following criteria: **1)** Germline variants failing any of the hard-filtering quality criteria were discarded, keeping only those that passed the three criteria considered; **2)** As an additional layer of confidence, we excluded from our cohort all the germline variants flagged in gnomAD v2.1.1 as failing at least one of their rigorous quality control filtering criteria (VCFs containing variants from 125,748 exomes and 15,708 genomes aligned to the reference genome hg19 were downloaded from: https://gnomad.broadinstitute.org/downloads#v2); **3)** Variants having at least 4 alternate reads were retained; **4)** Variants having at least 20 reads of total depth were kept; **5)** Variants with a Variant Allele Fraction (VAF) of at least 0.1 were kept; **6)** Variants with exonic regions and those in splice sites were retained; **7)** Missense, nonsense, frameshift insertions/deletions (INDELs), non-frameshift INDELs, start-loss, stop-loss, and splice site donor or acceptor variants were kept; **8)** Variants were retained if they had a population allele frequency lower than 0.01 in both ExAC and gnomAD (v2.1.1), or lower than 0.01 in one database but absent in the other, or not present in both; **9)** Variants classified in ClinVar as pathogenic or likely pathogenic were kept; **10)** For those annotated in ClinVar as conflicting, Variants of Uncertain Significance (VUS), or lacking a classification, only missense, nonsense, splicing, and frameshift INDELs were considered and further evaluated with *in silico* predictions. We ascribed as likely pathogenic those meeting the following criteria (and therefore retained them): **A)** Missense variants: predicted as deleterious/damaging by at least 4 of 6 tools (MutationTaster[14], PROVEAN[15], CADD[16], M-CAP[17], METASVM[18], FATHMM-XF[19], Mutation Assessor[20]), with the additional requirement of both CADD > 20 and an AlphaMissense[21] pathogenic score (>0.564). In addition, missense variants predicted as disease-causing exclusively by MutationTaster, provided they also accounted with both CADD > 20 and an AlphaMissense pathogenic score (>0.564) were also ascribed as likely pathogenic. **B)** Truncating-predicted variants: splice-site donor/acceptor variants with both SpliceAI[22] delta > 0.5 and dbscSNV > 0.6 scores were retained. Nonsense and frameshift INDELs were also kept with no additional *in silico* prediction filters; **11)** To reduce potential residual noise, we excluded genes in which 3 or more variants were observed in the same gene of the same patient, which may indicate possible technical artifacts or highly polymorphic genes rather than true biological events; **12)** We also excluded the olfactory receptor genes based on the findings of a previous study in the field, which demonstrated that olfactory receptor genes tend to have high mutation rates but low expression levels, which makes them prone to false-positive associations in cancer analyses[23]. In addition to this point, though with some exceptions recently found, it has been observed that most olfactory receptor genes are intronless[24], with their coding sequence contained in a single exon. Consistent with this, most variants identified in olfactory receptor genes in our cohort were located in exon 1. Based on these observations, we discarded these genes from downstream association analyses. After applying all the filtering criteria described above, the resulting final list of germline variants were considered as GPVs.

***Second hit analysis***

We determined the somatic second hit status of each GPV per patient in the cohort, either as **1)** somatic point mutations/small INDELs or **2)** as Loss of Heterozygosity (LOH) through the loss of the wild type allele. For **1)**, we considered the following criteria: **A)** Somatic mutations with a ClinVar classification (if any) of pathogenic or likely pathogenic; **B)** Somatic mutations with either a truncating-predicted effect, such as nonsense, splice site and frameshift INDELs, or missense if it was classified as damaging or deleterious by both PolyPhen and SIFT, based on the annotations provided in the MAF file of the MC3 project. For **2)** a GPV was considered to have LOH in the tumour if the second allele was lost by: **A)** Copy-Neutral LOH (CN-LOH) when the integer total copy number is 2 and the minor allele is 0; **B)** Duplication-LOH (DUP-LOH) when the integer total copy number is >2 and the minor allele is 0; **C)** Hemizygous-LOH (HEM-LOH) when the integer total copy number is 1 and the minor allele is 0. Somatic homozygous deletions were considered as the somatic full loss of both alleles. From the 824 cases in the cohort, 30 had either missing or unavailable CNV data and were excluded. This resulted in a total final cohort of 794 patients that were included in all subsequent analyses.

***Sig3 analysis***

We analyzed Sig3 presence for each individual patient's tumour (n = 794) using SigMA[25] (https://github.com/parklab/SigMA/wiki), a computational algorithm with a unique machine-learning boosted capacity to accurately detect Sig3 in samples with low mutation count. Utilizing as input the MAF file containing the somatic mutations of the MC3 project specifically from the breast cohort, a Sig3 score (Signature_3_mva) calculated by the gradient boosting classifier of the algorithm was obtained for each tumour.

**Supplementary Figure 1. Germline and somatic mutational landscape of Sig3+ patients (n = 141).**

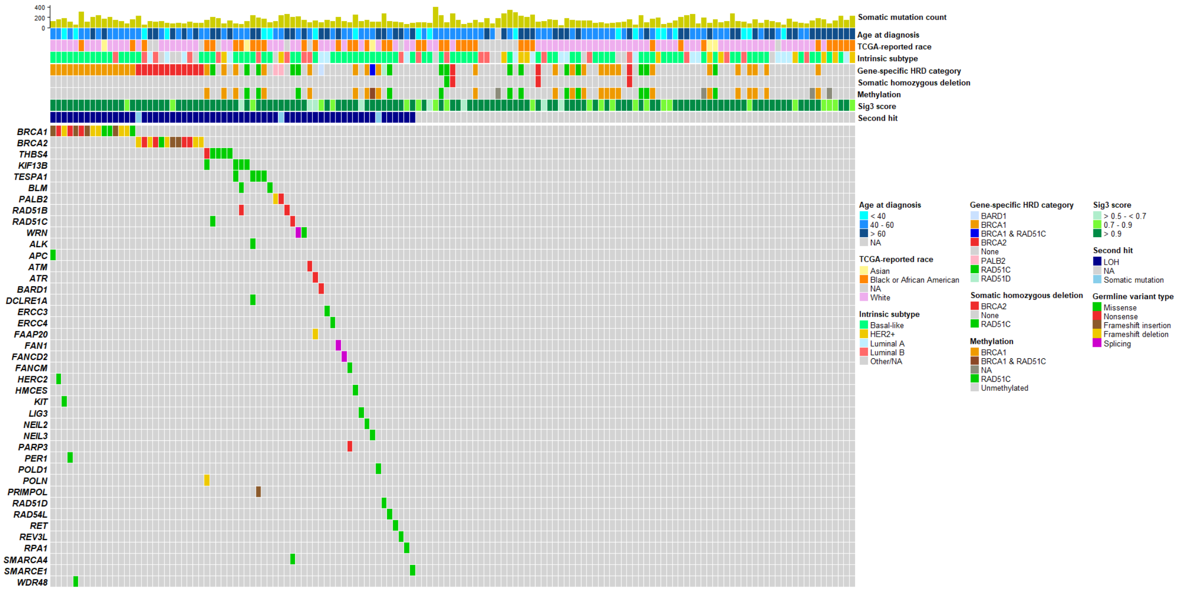


All the GPVs illustrated harbored a second hit in the tumour, as illustrated in the track "Second hit". Abbreviations: TCGA-BRCA: Breast cancer exome dataset of The Cancer Genome Atlas. HRD: Homologous Recombination Repair Deficiency. Sig3: Signature 3. NA: Not Applicable. HER2+: Human Epidermal Growth Factor Receptor 2 positive (HER2+). LOH: Loss of Heterozygosity.

**Supplementary Table 1. Datasets utilized in our study.**

| Required datasets | Source | Availability |
|---|---|---|
| Germline:<br>Exome variants | GDC platform | Unavailable (processed through in-house pipeline) |
| Tumor:<br>Exome variants<br>CNVs<br>Promoter methylation (*BRCA1/RAD51C*) | <br>GDC platform<br>GDC platform<br>DOI: 10.1038/ng.3934 | <br>Publicly available<br>Publicly available<br>Upon request |
| Clinical:<br>Intrinsic subtype<br>Age at diagnosis<br>TCGA-reported race | cBioPortal | Publicly available |

Summary of the germline, tumour and clinical datasets analyzed from TCGA-BRCA, including their corresponding sources and availabilities. Abbreviations: GDC: The Genomic Data Commons. TCGA: The Cancer Genome Atlas. CNVs: Copy Number Variants.

**Supplementary Data 1. Framework gene panel.** The panel was curated in-house by merging clinical-grade panels for cancer susceptibility offered by different diagnostic laboratories, as well as genes evaluated in two of the largest case-control studies in hereditary breast cancer[26,27], a comprehensive list of DNA-repair genes, and the genes that reached statistical significance ($p < 0.05$) in the exome-wide unbiased association analyses. The first sheet summarizes the sources of each gene list, which includes the gene panel name, size, and source links. The subsequent sheets provide the full gene panels listed in the first sheet.

**Supplementary Data 2. Exome-wide association with Sig3 of GPVs with somatic second hits, which included patients carrying GPVs with second hits in the well-known HRD genes *BARD1, BRCA1/2, PALB2,* and *RAD51C/D.*** Barnard's exact test, two-sided. ORs, CI, p-values, and adjusted p-values are listed for each gene tested, including the number of carriers of GPVs with second hits per gene. The log2 (ORs) and -log10 (adjusted p-values) values correspond to the volcano plot illustrated in Figure 2. Abbreviations: ORs: Odds ratios. CI: Confidence Intervals. Sig3+: Signature 3 positive. Sig3-: Signature 3 negative.

**Supplementary Data 3. Exome-wide association with Sig3 of GPVs with somatic second hits, which excluded patients carrying GPVs with second hits in the well-known HRD genes *BARD1, BRCA1/2, PALB2,* and *RAD51C/D.*** Barnard's exact test, two-sided. ORs, CI, p-values, and adjusted p-values are listed for each gene tested, including the number of carriers of GPVs with second hits per gene. The log2 (ORs) and -log10 (adjusted p-values) values correspond to the volcano plot illustrated in Figure 3. Abbreviations: ORs: Odds ratios. CI: Confidence Intervals. Sig3+: Signature 3 positive. Sig3-: Signature 3 negative.

**Supplementary Data 4. Germline, tumour and clinical data of Sig3+ patients (n = 11/141) with BLBC identified through the clinico-genomics approach in the TCGA-BRCA cohort.** Only GPVs with evidence of a somatic second hit are included. No HRD-related genetic events (GPVs with second hits and somatic homozygous deletions in *BARD1, BRCA1/2, PALB2,* and *PALB2*, as well as somatic promoter methylation in *BRCA1/RAD51C*) were observed in such patients. Abbreviations: TCGA: The Cancer

Genome Atlas. Sig3: Signature 3. BLBC: Basal-like Breast Cancer. LOH: Loss of Heterozygosity.

**Supplementary Data 5. Germline, tumour and clinical data of both Sig3+ and Sig3- breast cancer patients (n = 8/794) carrying GPVs with second hits in the gene *FANCM*.** In such patients, no HRD-related genetic events (GPVs with second hits and somatic homozygous deletions in *BARD1, BRCA1/2, PALB2,* and *RAD51C/D,* as well as somatic promoter methylation in *BRCA1/RAD51C*) were identified. Abbreviations: TCGA: The Cancer Genome Atlas. Sig3: Signature 3. BLBC: Basal-like Breast Cancer. LOH: Loss of Heterozygosity.
